# Sustainability Indexes as Possible Predictors of Excess Mortality across OECD Countries during the Covid-19 Pandemic

**DOI:** 10.1101/2023.06.09.23291035

**Authors:** Lee Liu

## Abstract

This study considered the Sustainable Development Goals (SDG) Index, Human Development Index (HDI), and Environmental Performance Index (EPI) as sustainability indexes and explored their potential effectiveness as predictors of Covid-19 excess mortality among countries in the Organization for Economic Cooperation and Development (OECD). The results indicated strong negative correlations between each index and Covid-19 excess mortality. Each of the indexes was able to consistently predict variations in excess mortality in the regression models. Such findings contribute to the current discussion on what lessons we should take away from the Covid-19 pandemic, specifically whether sustainability indexes may be useful in assessing Covid-19 outcomes. It is valuable to further examine the role sustainable policies and practices can play in preparing for future crises, as it has been demonstrated that social, economic, and environmental factors influenced the ability of a country to mitigate the impact of Covid-19. Overall, the most effective way for a country to both prevent and prepare for the next crisis is to improve health, education, and standard of living for its people, protect the environment, and achieve sustainability.

## Introduction

The Organization for Economic Cooperation and Development (OECD) has published its report on lessons we should learn from the Covid-19 pandemic (OECD, 2023). The main takeaways include strengthening health systems that are underprepared and understaffed due to underinvestment, addressing socio-economic problems, such as inequality, poverty, and unemployment, improving governance and trust, specifically regarding data collection and sharing, and implementing greater coordination and cooperation (OECD, 2023). Similar lessons have been discussed in other publications (Covid Crisis Group, 2023; Bollyky, et al., 2023; Berchet et al., 2023; Sachs JD, et al., 2022; WHO-SEARO, 2022; Gupta, 2021). The OECD calls for investing in health system resilience to prepare for the next crisis (OECD, 2023). It also recognizes that health systems are a part of the larger social systems as it was successful in implementing “whole-of-society responses” to the pandemic (OECD, 2023).

Although the issues being raised are all tied to sustainability, a sustainability perspective is frequently missing from these conversations. This paper intends to address this gap by examining whether sustainability indexes could be used as predictors of Covid-19 pandemic outcomes. Specifically, the research explores variations in excess mortality across OECD countries during the pandemic with attention to three indexes: the Sustainable Development Goals (SDG) Index, Human Development Index (HDI), and Environmental Performance Index (EPI).

Excess mortality reflects the “true death toll of COVID-19 (WHO, 2023)”. By the end of 2021, global excess deaths were estimated to be 14.83 million, although only 5.42 million deaths were attributed to Covid-19 (Msemburi et al., 2023). The SDG Index, HDI, and EPI should be able to provide insight into the overall development and preparedness of OECD countries in addressing the Covid-19 pandemic. Though not specifically designed as predictors of excess deaths due to Covid-19, they can still provide context regarding the social, economic, and environmental circumstances of a country that could have influenced its ability to manage and mitigate the impact of the pandemic.

The paper uses a broad definition of sustainability to include indexes that measure only some components of sustainability. The SDG Index assesses countries’ progress towards achieving the United Nations’ 17 Sustainable Development Goals (SDGs), which include goals related to poverty eradication, healthcare, education, equality, and sustainable development (Sachs J et al., 2022). The index can provide an overall picture of a country’s development and resilience. Countries with higher SDG Index scores generally have stronger social systems and healthcare infrastructure, which could potentially contribute to better handling of the pandemic and lower excess deaths. The HDI measures a country’s average achievements in improving human well-being and quality of life (health, education, and living standards) (UNDP, 2022), which are important components of sustainability and associated with some SDGs (Neumayer 2010; Conceição 2019). The EPI ranks countries based on their environmental performance and sustainability. It evaluates various factors such as air and water quality, biodiversity, climate change mitigation, and environmental health (Wolf et al., 2022). SDG index scores have been reported to be associated with Covid-19 mortality among Asian countries (Zhou and Puthenkalam, 2022) and among US states (Liu, 2023). In both cases, places with higher SDG index scores tend to experience lower mortality. It has been reported that places that made progress in achieving SDGs before Covid-19 did better in pandemic mitigation, while places lacking progress in achieving the SDGs experienced more severe impacts of the pandemic (UN/DESA, 2020). Reports on the effects of HDI scores on Covid-19 outcomes have been mixed (Mirahmadizadeh et al., 2022; Heo et al., 2022). However, there is a lack of research in the relationship between EPI rank and Covid-19 mortality or the effects of the three indexes on Covid-19 mortality among the OECD countries.

## Data and Methods

The 2020–2021 excess death rates among OECD countries were obtained from the OECD report *Ready for the Next Crisis? Investing in Health System Resilience* (OECD, 2023). They were the percentage changes in annual all-cause mortality from 2015-2019 to 2020-2021 (OECD, 2023). Data were unavailable for three of the 38 OECD member countries: Costa Rica, Ireland, and Türkiye.

The HDI scores (from 0 to 100) of the OECD countries were from the UNDP GLOBAL 2021/22 Human Development Report (UNDP, 2022). The EPI scores (from 0 to 100) were from the *2022 Environmental Performance Index* (Wolf et al., 2022), The SDG index scores (from 0 to 100) were from the Sustainable Development Report 2022 (Sachs J et al., 2022).

To avoid using cofounding variables that might bias the results, this study looked for other variables that have been reported to affect mortality but are not directly correlated to the three indexes. It found data available for vaccination rates and emotional stress as appropriate variables to be included in the analyses. OECD (2023) provided data on emotional stress across all OECD countries except for Belgium, Chile, Israel, and Luxembourg. Emotional stress was expressed as the percentage of people expressing sadness in the 2020 *Gallup Global Emotions Survey*. Vaccination rates were based on percentage of population fully vaccinated by the end of 2021 according to Covid-19 data from Our World in Data (2023). The source did not have data on Luxembourg, Switzerland, or Türkiye.

Demographic characteristics have been found to influence Covid-19 outcomes (ONS, 2022; Chang, 2022). Thus, this research used two demographic features with data from OECD Data (2023). One was the elderly population ratio (people aged 65 and over as percentage of total population) with data missing for Colombia and Costa Rica. The other was the elderly dependency ratio defined as the number of elderly people compared to the working age population at ages 20 to 64. Such data were unavailable for Chile, Costa Rica, Iceland, and Ireland. Gini coefficient (as a measurement of income inequality) data were also from OECD Data (2023), with data missing for Costa Rica and Ireland. Data on population density (people per sq. km of land area) was obtained from the World Bank (2023).

The analyses on the relationship between the excess deaths and possible explanatory variables were carried out using the SPSS software. Pearson correlation analyses were performed on all variables. They were followed with linear regression analyses to further investigate the possible explanatory factors of the excess death rates with special attention to the HDI, EPI, and SDG indexes. To minimize cofounding effects, a regression model used only one of the three indexes. Additionally, dependency ratio and elderly population ratio were not used in the same model, and Gini coefficient was not included in the same model as SDG index, which emphasizes equality.

## Results

The descriptive statistics show the large differences in excess all-cause mortality among the 35 OECD countries for which data were available (Table 1). The lowest excess death rate was found in Norway, which was only 1.2% above normal. The highest rates were 48% in Colombia and 55.9% in Mexico. These two countries were ranked the lowest in both the SDG index and HDI. Denmark, Finland, and Sweden scored the highest in SDG index, while Switzerland and Norway had the highest HDI scores. In terms of EPI, Colombia and Mexico were among the lowest. The highest EPIs were found in Denmark, the UK, Finland, and Sweden. Vaccination rates were the lowest in Slovak Republic and Poland and highest in Chile, Portugal, South Korea, and Japan. People’s mental stress was lowest in Japan, followed by Iceland and Finland. The highest stress scores were found in Türkiye, Colombia, and Mexico. The lowest population densities were found in Australia, Iceland, and Canada while the highest were in South Korea, the Netherlands, and Israel. The old age dependency ratios were the lowest in Latvia and Lithuania and highest in Japan and Finland. Mexico and Colombia were the lowest in terms of aging while Japan and Italy were the highest. The lowest Gini coefficients were found in Slovak Republic and Slovenia while the highest were in Colombia and Mexico.

**Table 1.** Descriptive statistics.

|  | N | Minimum | Maximum | Mean | Std. Deviation |
| --- | --- | --- | --- | --- | --- |
| Excess death rate (%) | 35 | 1.2 | 55.9 | 13.58 | 11.34 |
| SDG Index score (0–100) | 38 | 70.1 | 86.5 | 78.69 | 3.84 |
| HDI score (0–100) | 38 | 76.7 | 96.2 | 90.13 | 4.83 |
| EPI score (0–100) | 38 | 26.3 | 77.9 | 58.01 | 10.42 |
| Vaccination rate (%) | 36 | 43.56 | 84.41 | 69.22 | 9.32 |
| Emotional stress (%) | 34 | 11.2 | 49.5 | 22.23 | 7.45 |
| Population density | 38 | 3 | 531 | 139.11 | 140.51 |
| Dependency ratio (%) | 36 | 3.1 | 53.4 | 30.07 | 9.73 |
| Elderly population (%) | 34 | 7.86 | 28.86 | 18.71 | 4.23 |
| Gini coefficient | 36 | 0.222 | 0.487 | 0.31 | 0.05 |

All predictor variables are significantly correlated to excess death rate and in the expected direction according to prior knowledge except for population density (Table 2). Thus, population density was excluded in the linear regression analyses. The regressions examined how excess death rates among the OECD countries were affected by the three indexes, vaccination, mental stress, elderly ratio, old age dependency ratio, and Gini coefficient.

**Table 2.** Pearson correlation.

| Predictors | Excess death rate (%) |  |  |
| --- | --- | --- | --- |
|  | Coefficients | Sig. (2-tailed) | N |
| SDG Index score (0–100) | −0.654** | 0 | 35 |
| HDI score (0–100) | −0.849** | 0 | 35 |
| EPI score (0–100) | −0.531** | 0.001 | 35 |
| Vaccination rate (%) | −0.565** | 0.001 | 33 |
| Emotional stress (%) | 0.631** | 0 | 31 |
| Population density | −0.135 | 0.438 | 35 |
| Dependency ratio | −0.349* | 0.043 | 34 |
| Elderly ratio | −0.619** | 0 | 33 |
| Gini coefficient | 0.578** | 0 | 34 |

\* indicates significance at 95%.
\*\* indicates significance at 99.9%.

The SDG index appears to be a significant explainer of the variations of excess death rate among the countries (Table 3). It was consistently the only explanatory factor with a significance of 95% in all six models. Vaccination rate was another key factor, followed by emotional stress. Elderly ratio was significant in only one model while dependency was insignificant in any of the models. The high R^2^ values suggest that the models were able to explain over 60% of the changes in the excess death rates in all regression models, except for Model C, which was over 45%.

**Table 3.**
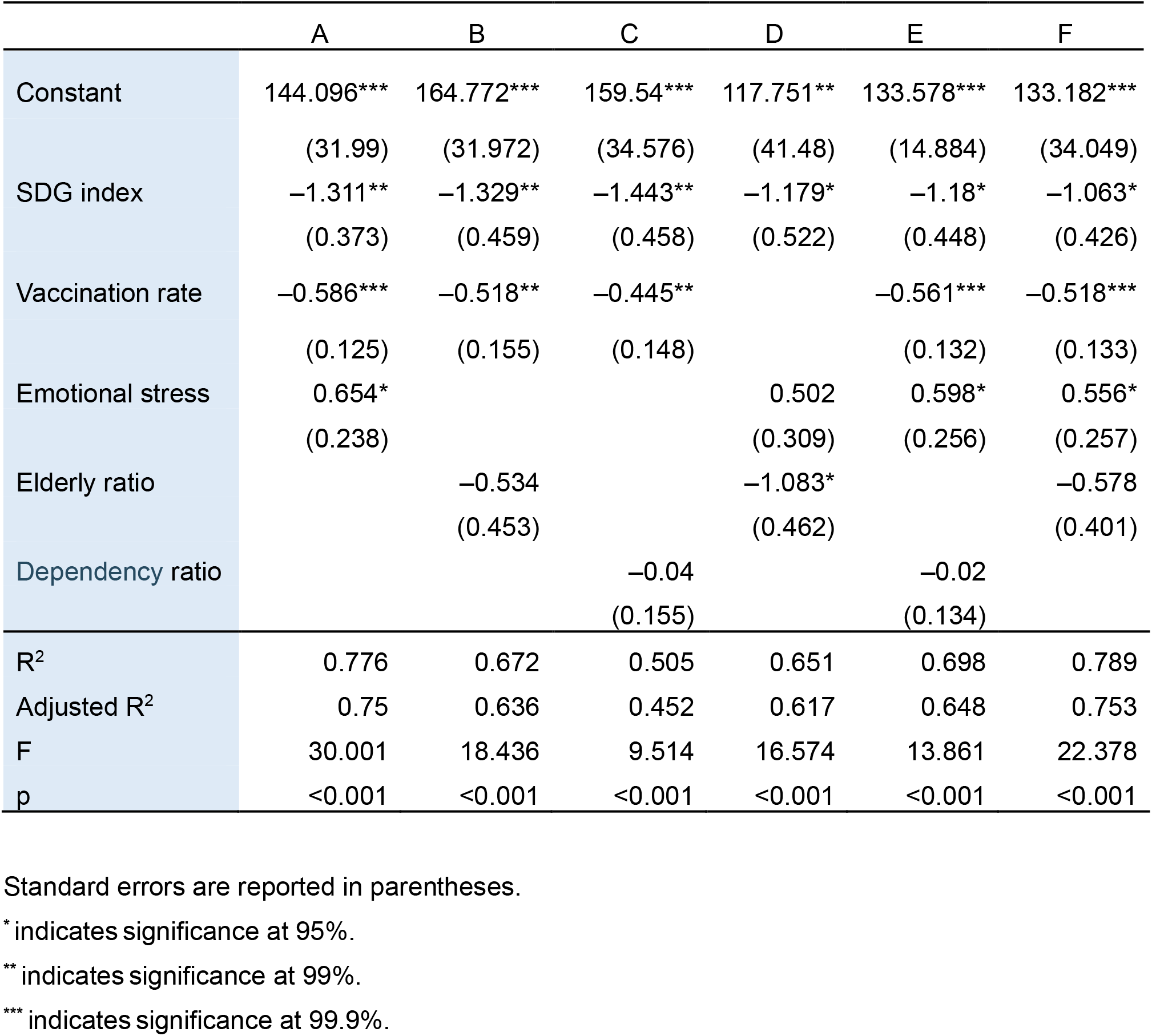
Regression results for Covid-19 associated excess death rates among OECD countries, 2020-2021, SDG index models.

The HDI was also consistently a key explanatory factor from Regressions A through F (Table 4). Vaccination rate was also a primary explanatory factor followed by emotional stress scores. Elderly ratio and Gini coefficient had a significant effect in only one of the six regressions, while dependency did not have any effects. All regressions were highly significant and with adjusted R^2^ over 0.8 in five of the six models, indicating that the models were able to explain over 80% of the variations in the excess death rates.

**Table 4.**
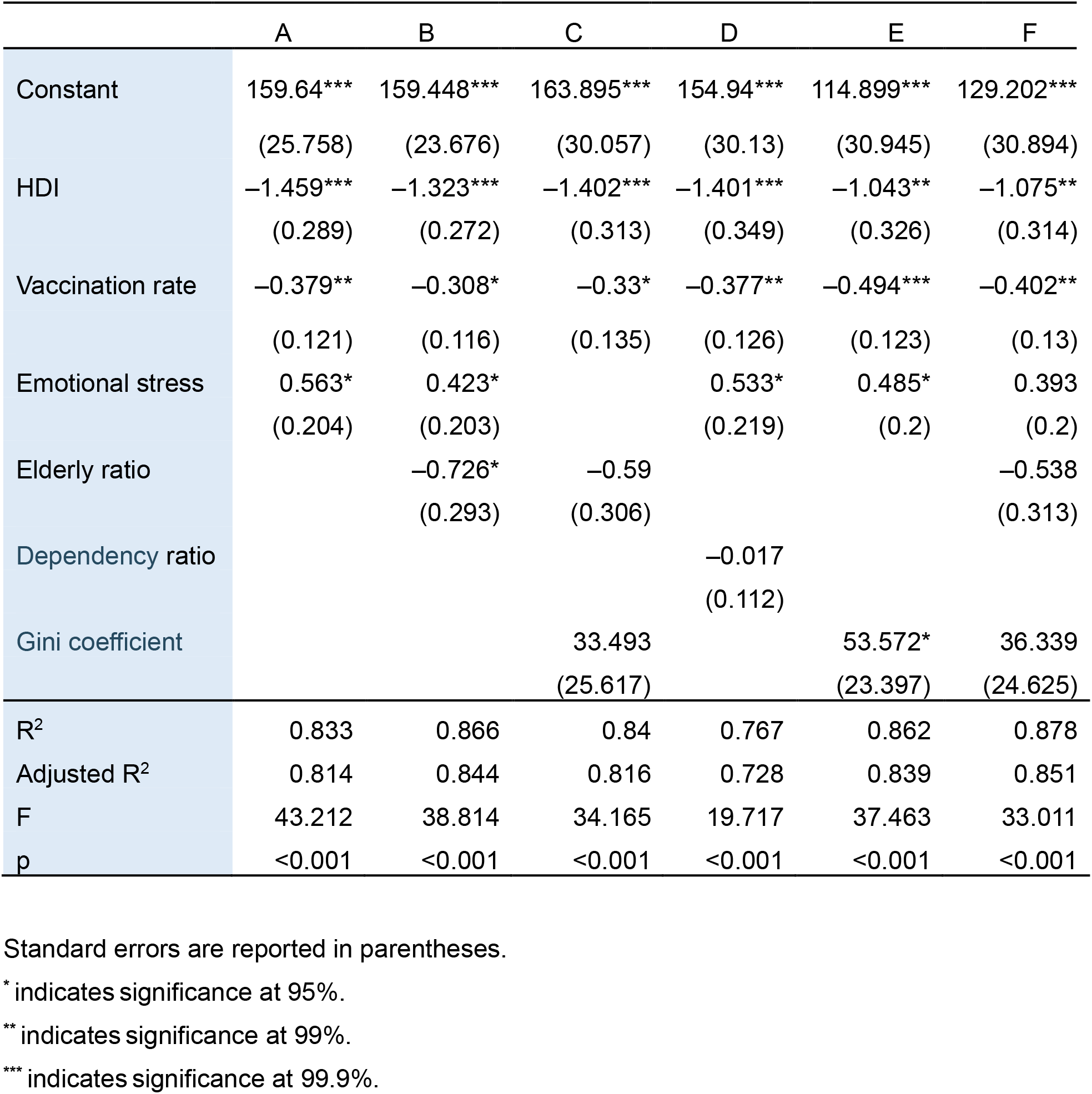
Regression results for Covid-19 associated excess death rates among OECD countries, 2020-2021, HDI models.

|  | A | B | C | D | E | F |
| --- | --- | --- | --- | --- | --- | --- |
| Constant | 159.64***<br>(25.758) | 159.448***<br>(23.676) | 163.895***<br>(30.057) | 154.94***<br>(30.13) | 114.899***<br>(30.945) | 129.202***<br>(30.894) |
| HDI | -1.459***<br>(0.289) | -1.323***<br>(0.272) | -1.402***<br>(0.313) | -1.401***<br>(0.349) | -1.043**<br>(0.326) | -1.075**<br>(0.314) |
| Vaccination rate | -0.379**<br>(0.121) | -0.308*<br>(0.116) | -0.33*<br>(0.135) | -0.377**<br>(0.126) | -0.494***<br>(0.123) | -0.402**<br>(0.13) |
| Emotional stress | 0.563*<br>(0.204) | 0.423*<br>(0.203) |  | 0.533*<br>(0.219) | 0.485*<br>(0.2) | 0.393<br>(0.2) |
| Elderly ratio |  | -0.726*<br>(0.293) | -0.59<br>(0.306) |  |  | -0.538<br>(0.313) |
| Dependency ratio |  |  |  | -0.017<br>(0.112) |  |  |
| Gini coefficient |  |  | 33.493<br>(25.617) |  | 53.572*<br>(23.397) | 36.339<br>(24.625) |
| $R^2$ | 0.833 | 0.866 | 0.84 | 0.767 | 0.862 | 0.878 |
| Adjusted $R^2$ | 0.814 | 0.844 | 0.816 | 0.728 | 0.839 | 0.851 |
| F | 43.212 | 38.814 | 34.165 | 19.717 | 37.463 | 33.011 |
| p | <0.001 | <0.001 | <0.001 | <0.001 | <0.001 | <0.001 |

Standard errors are reported in parentheses.
\* indicates significance at 95%.
\*\* indicates significance at 99%.
\*\*\* indicates significance at 99.9%.

Similar to the SDG index and HDI, the EPI was also a key predictor with consistent effects in all models (Table 5). Vaccination rate was also a key factor in predicting excess mortality in all models. Emotional stress was able to produce an effect in three of the six models. Elderly ratio and Gini coefficient had a significant effect in one of the models. Dependency’s effects were insignificant. All models achieved a significance at 99.9%, with adjusted R^2^ values over 0.6 in five of the six models.

**Table 5.**
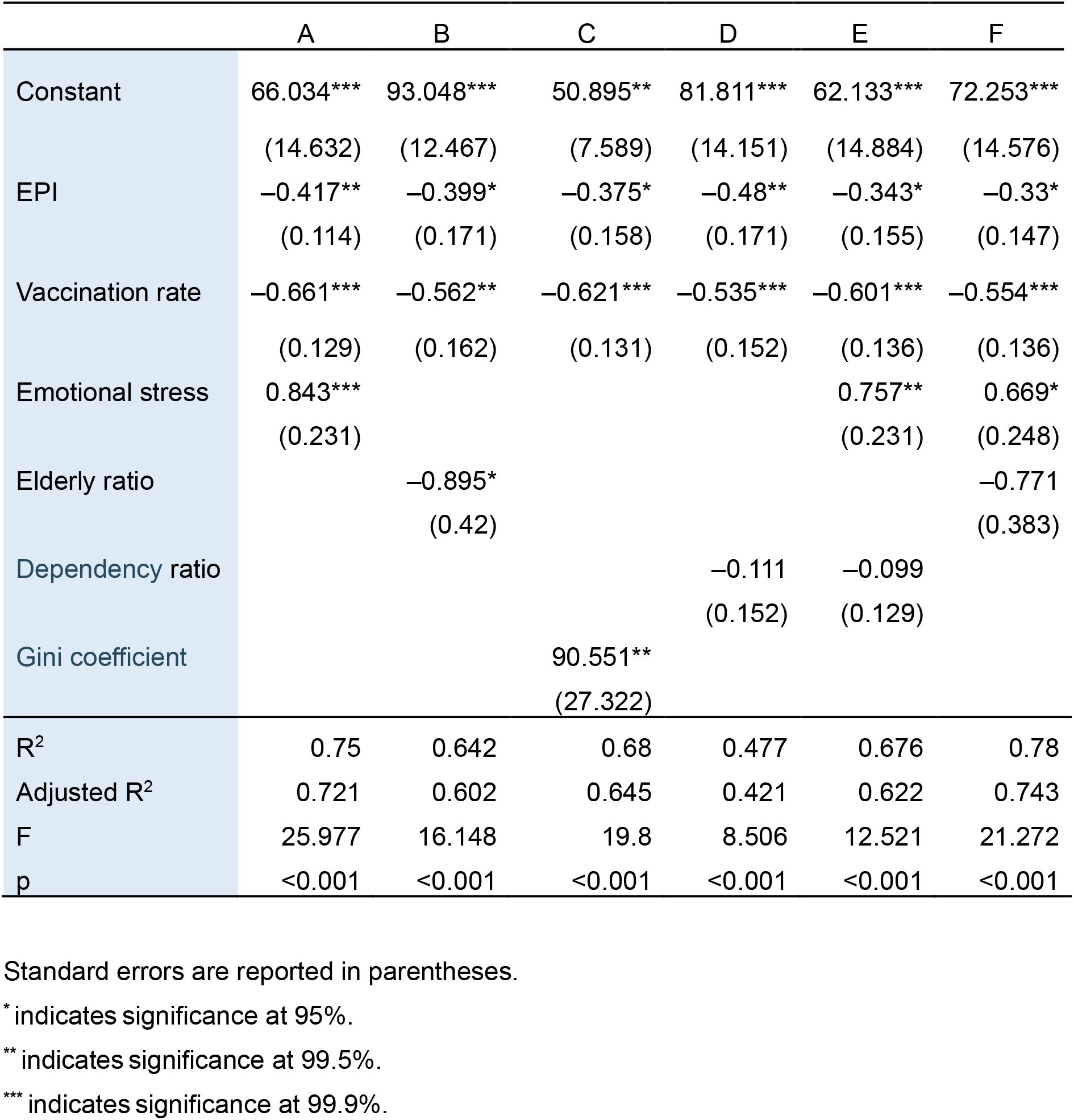
Regression results for Covid-19 associated excess death rates among OECD countries, 2020-2021, EPI models.

|  | A | B | C | D | E | F |
| --- | --- | --- | --- | --- | --- | --- |
| Constant | 66.034***<br>(14.632) | 93.048***<br>(12.467) | 50.895**<br>(7.589) | 81.811***<br>(14.151) | 62.133***<br>(14.884) | 72.253***<br>(14.576) |
| EPI | -0.417**<br>(0.114) | -0.399*<br>(0.171) | -0.375*<br>(0.158) | -0.48**<br>(0.171) | -0.343*<br>(0.155) | -0.33*<br>(0.147) |
| Vaccination rate | -0.661***<br>(0.129) | -0.562**<br>(0.162) | -0.621***<br>(0.131) | -0.535***<br>(0.152) | -0.601***<br>(0.136) | -0.554***<br>(0.136) |
| Emotional stress | 0.843***<br>(0.231) |  |  |  | 0.757**<br>(0.231) | 0.669*<br>(0.248) |
| Elderly ratio |  | -0.895*<br>(0.42) |  |  |  | -0.771<br>(0.383) |
| Dependency ratio |  |  |  | -0.111<br>(0.152) | -0.099<br>(0.129) |  |
| Gini coefficient |  |  | 90.551**<br>(27.322) |  |  |  |
| $R^2$ | 0.75 | 0.642 | 0.68 | 0.477 | 0.676 | 0.78 |
| Adjusted $R^2$ | 0.721 | 0.602 | 0.645 | 0.421 | 0.622 | 0.743 |
| F | 25.977 | 16.148 | 19.8 | 8.506 | 12.521 | 21.272 |
| p | <0.001 | <0.001 | <0.001 | <0.001 | <0.001 | <0.001 |

Standard errors are reported in parentheses.
\* indicates significance at 95%.
\*\* indicates significance at 99.5%.
\*\*\* indicates significance at 99.9%.

## Discussion

The results indicate that all three indexes could be possible predictors of excess mortality as they significantly affect the excess mortality in the OECD countries. Their effects were consistent with different models controlling for other possible predictors such as vaccination rates, emotional stress, population characteristics, and Gini coefficient. Vaccination rates also proved to be a strong predictor, followed by emotional stress. However, the effects of elderly ratio, old age dependency ratio, and Gini coefficient were often insignificant when used with any of the three indexes in a regression model. Population density did not significantly correlate to the death rates.

It was not surprising that the three indexes were able to predict the excess mortality among the OECD countries, as a country’s achievement in sustainability would enable more effective pandemic mitigation. Furthermore, the main lessons described in the OECD report (OECD, 2023) are ultimately lessons in sustainable development as discussed below.

1. The problem of underprepared and understaffed health systems could have been eased by achieving SDG 3, which requires countries to strengthen their healthcare system capacity to promote health and well-being for everyone (UN, 2015). Quality healthcare and universal health coverage are also necessary components of SDG 3 (UN, 2015), which would have helped improve health system resilience. Higher HDI scores typically indicate better healthcare systems and higher living standards, which can play a role in managing the impact of a pandemic. Countries with higher HDI scores might have more robust healthcare infrastructure, better access to medical resources, and stronger public health systems, which could potentially help reduce excess deaths during the Covid-19 pandemic. While the EPI primarily focuses on environmental indicators, a country’s environmental performance can indirectly influence its ability to address public health crises. For example, countries with cleaner air and water might have populations with better respiratory health, which could potentially reduce the severity of Covid-19 outcomes. People living in countries with better environmental conditions may also experience less emotional stress during the pandemic, as it has been reported that higher exposure to green spaces was associated with lower Covid-19 deaths among US counties (Yang et al., 2022).
2. Socio-economic problems such as inequality, poverty, and unemployment could have been managed by achieving a number of SDGs: SDG 1 for poverty elimination, SDG 3 for health equity, SDG 10 for reducing income inequalities, and SDG 11 for developing inclusive cities (UN, 2015). Similarly, countries achieving higher HDIs tend to have less poverty and better employment opportunities.
3. The challenges of governance and trust during the pandemic would have been better met if SDG 16 had been achieved, which calls for “effective, accountable and transparent institutions” (UN, 2015). Countries with higher HDI and EPI scores tend to have stronger governance and public trust when dealing with crises, which can aid in managing a pandemic.
4. The three indexes also promote data collection and sharing, and coordination and cooperation. The SDGs emphasize collaboration among governments, NGOs, civil society, and the private sector (UN, 2015). Cooperation and coordination require data sharing to make effective collective efforts in mitigation. They are the required targets in SDG 11 for inclusive societies and SDG 16 for strong institutions (UN, 2015). Achieving higher HDI and EPI scores also require strong institutions that practice coordination and cooperation.

Ultimately, the findings contribute to the debates over lessons we should learn from the pandemic (OECD, 2023; Covid Crisis Group, 2023; Bollyky, et al., 2023; Berchet et al., 2023; Sachs JD, et al., 2022; WHO-SEARO, 2022; Gupta, 2021) and the ability of SDG index and HDI to predict Covid-19 outcomes (Zhou and Puthenkalam, 2022; Liu, 2023; UN/DESA 2020; Mirahmadizadeh et al., 2022; Heo et al., 2022).

## Limitations

Several limitations must be noted. First, the data used in the statistical analyses have limitations. Excess mortality data were missing for three of the OECD countries. Missing data were also an issue with vaccination rates and population features. The three indexes are composite indicators, which must join many component indicators using arbitrary weighting (Conceição, 2019). The final scores have limitations when they are used for comparing countries. For example, the SDG index scores are based on 232 indicators. Some indicators had to be measured using dated data due to the unavailability of more recent data and data for some indicators were unavailable (Lynch and Sachs, 2021). Second, there are different methods of estimation for excess death rates, and the rates may be different due to the methods used. Third, there are possibly other significant predictors of excess death rates for which data were unavailable and thus excluded from the analyses. Last, a strong correlation between the dependent and independent variables does not necessarily mean cause and effect. Other factors or explanations may exist that contribute to the observed relationship, as Covid-19 outcomes are influenced by a complex interplay of factors. While indexes like the SDG Index, HDI, and EPI can provide insights into a country’s overall development and resilience, they should be considered as just some of the many factors that could potentially influence excess deaths during the pandemic among OECD countries. Thus, caution is required when interpreting the findings.

## Conclusions

The findings indicate that sustainability indexes (SDG index, HDI, and EPI) could be powerful predictors of the changes in excess death rates during the Covid-19 pandemic among OECD countries. Such findings should be able to contribute to the current debates over the lessons countries should learn from the pandemic and the usefulness of sustainability indexes in predicting Covid-19 outcomes. The indexes were able to act as predictors because they provided context regarding a country’s social, economic, and environmental factors that could determine its ability to manage and mitigate the impact of the pandemic. Lessons from the pandemic, as recognized by the OECD or elsewhere, were all related to sustainability issues. Sustainability is probably the best investment in health system resilience to prepare for the next crisis. The next crises could be another pandemic, climate change, environmental emergencies, armed conflict, or social unrest. The most essential preparation that countries need to make for these crises is to improve people’s health, education, and standard of living, protect the environment, and achieve sustainability.

## Data Availability

The research was based on publicly available data, as stated in the Data and Methods section.

## Acknowledgements

The author thanks Tiffany Liu for editing the paper.

